# CSF p-tau181 Increases in Patients with Neuronal Intranuclear Inclusion Disease without Amyloid Burden

**DOI:** 10.1101/2022.06.09.22275750

**Authors:** Masanori Kurihara, Hiroki Komatsu, Renpei Sengoku, Mari Shibukawa, Satoru Morimoto, Tomoyasu Matsubara, Akira Arakawa, Makoto Orita, Kenji Ishibashi, Akihiko Mitsutake, Shota Shibata, Hiroyuki Ishiura, Kaori Adachi, Kensuke Ohse, Keiko Hatano, Ryoko Ihara, Mana Higashihara, Yasushi Nishina, Aya Midori Tokumaru, Kenji Ishii, Yuko Saito, Shigeo Murayama, Kazutomi Kanemaru, Atsushi Iwata

## Abstract

**Background and Objectives:** CSF tau phosphorylated at threonine 181 (p-tau181) is a widely used biomarker for Alzheimer’s disease (AD) and has recently been regarded to reflect amyloid-beta (Aβ) and/or p-tau deposition in the AD brain. Although it is important to know how this biomarker reacts in other neurocognitive diseases, CSF p-tau181 in patients with neuronal intranuclear inclusion disease (NIID) has not been studied.

**Methods:** CSF concentrations of p-tau181, total tau, amyloid-beta 1-42 (Aβ42), monoamine metabolites homovanillic acid (HVA), and 5-hydroxyindole acetic acid (5-HIAA) were compared between 12 patients with NIID, 120 patients with symptomatic AD biologically confirmed based on CSF biomarker profiles, and patients clinically diagnosed with other neurocognitive disorders (dementia with Lewy bodies [DLB], 24; frontotemporal dementia [FTD], 13; progressive supranuclear palsy [PSP], 21; and corticobasal syndrome [CBS], 13). Amyloid PET using Pittsburgh compound B (PiB) was performed in six NIID patients.

**Results:** CSF p-tau181 concentration was significantly higher in NIID (72.7 ± 24.8 pg/mL) compared to DLB, PSP, and CBS and was comparable between NIID and AD. CSF p-tau181 was above the cutoff value (50.0 pg/mL) in 11 of 12 NIID patients (91.7%). Within these patients, only two patients showed decreased CSF Aβ42, and these patients showed negative or mild local accumulation in PiB PET, respectively. PiB PET scans were negative in the remaining 4 patients tested. CSF HVA and 5-HIAA concentrations were significantly higher in patients with NIID compared to disease controls.

**Discussion:** CSF p-tau181 was increased in patients with NIID without amyloid accumulation. Although the deposition of p-tau has not been reported in NIID brains, molecular mechanism of tau phosphorylation or secretion of p-tau may be altered in NIID.

## Introduction

CSF tau phosphorylated at threonine 181 (p-tau181) is a widely used biomarker for Alzheimer’s disease (AD), the most common cause of dementia around the globe, and is also used in recent research criteria to classify patients in the AD continuum.^1^ AD is characterized by the presence of extracellular amyloid-beta (Aβ) plaque and hyperphosphorylated tau in neurofibrillary tangles in the brain; in the CSF, it is known that decreased Aβ 1-42 (Aβ42) (or Aβ42/Aβ40 ratio), increased p-tau181, and neurodegeneration occur in a sequential manner.^2,3^ Although there are other neurodegenerative diseases with p-tau deposition in the brain, consistently increased CSF p-tau181 is not observed in other neurodegenerative diseases and is considered to be specific to AD.^1,4,5^ Although it has recently been recognized that increased CSF p-tau181 reflects Aβ and/or p-tau deposition in the brain, the precise mechanism behind increased CSF p-tau181 remains unknown.^6^

Homovanillic acid (HVA) and 5-hydroxyindole acetic acid (5-HIAA) are the major metabolites of dopamine and serotonin, respectively. CSF concentrations of HVA and 5-HIAA have long been investigated in neurological and psychiatric diseases and are reported to be lower in several diseases, including Parkinson’s disease (PD) and dementia with Lewy bodies (DLB), likely reflecting reduced levels of dopamine and serotonin in the brain.^7–9^

Although it is also important to know how these biomarkers are changed in other neurodegenerative diseases, it is noteworthy that CSF biomarkers in patients with neuronal intranuclear inclusion disease (NIID) have yet to be thoroughly investigated. NIID is a neurodegenerative disease characterized by eosinophilic hyaline intranuclear inclusions in neurons, glial cells, and other somatic cells.^10,11^ Patients with NIID present with dementia, neuropathy, parkinsonism, or other various symptoms, and the number of reported patients presenting with dementia have significantly increased following the discovery of the diagnostic value of skin and other tissues biopsies.^10,12,13^ Although CGG repeat expansions in *NOTCH2NLC* have been identified as the cause of the disease in Asian patients with NIID,^14,15^ the precise pathophysiology of this disease is still under investigation. The CSF profile of patients with NIID is poorly studied, and only a mild increase of total protein has been reported.^10^

Therefore, in this study, we aimed to investigate whether CSF p-tau181, HVA, and 5-HIAA are altered in patients with NIID to further understand both the characteristics of these CSF biomarkers and the pathophysiology of NIID.

## Methods

### Participants and Settings

We retrospectively reviewed all patients who underwent CSF biomarker testing from April 2012 to March 2022 at the Tokyo Metropolitan Geriatric Hospital. The study protocols were approved by the Institutional Review Board of the Tokyo Metropolitan Institute of Gerontology (approval No. 250217). The study protocol for genetic testing in neurodegenerative diseases was approved by the Institutional Review Board of the University of Tokyo (approval No. G1396). Patients were recruited at the Tokyo Metropolitan Geriatric Hospital, and written informed consent was obtained from all participants or their caregivers.

Patients with a final diagnosis of NIID, based on the characteristics of the brain MRI findings and neuropathological assessment of skin biopsy tissues,^10,12,16^ were included as patients with NIID (n = 12). Electron microscopical assessment was conducted to differentiate intranuclear inclusions from p62-positive promyelocytic leukemia (PML) nuclear bodies observed in the nuclei of normal controls, as previously described.^16^ Patients with symptomatic AD biologically confirmed based on CSF biomarker profiles fulfilling A+T+ by the NIA-AA research framework criteria were included as AD (n=120).^1^ Patients with clinical diagnoses of DLB fulfilling probable according to the 2017 consensus criteria (n = 24),^17^ frontotemporal dementia (FTD) fulfilling the 1998 consensus criteria (n = 13),^18^ progressive supranuclear palsy (PSP) fulfilling probable according to the 2017 MDS-PSP criteria (n = 21),^19^ and corticobasal syndrome (CBS) fulfilling the modified Cambridge criteria (n = 13)^20^ were included as non-AD disease controls.

### CSF Measurements

CSF was obtained by standard lumbar puncture procedures. The first tube was sent for cell counting and routine biochemical testing, and the following CSF samples were directly collected in polypropylene tubes. CSF concentrations of p-tau181, t-tau, and Aβ42 were measured by ELISA (Fujirebio Inc., Gent, Belgium), in accordance with the manufacturer’s protocol, as previously described.^21^ The institutional cut-off values of p-tau181, 50.0 pg/mL; t-tau, 300 pg/mL; and Aβ42, 500 pg/mL were predetermined based on the results obtained from patients with neuropathological diagnosis at autopsy.^9^ Based on recent research criteria for AD, patients were classified as A-T-, A+T-, A-T+, or A+T+, according to the presence of abnormal CSF Aβ42 (< 500 pg/mL) and p-tau181 (> 50.0 pg/mL) results.^1^ CSF concentrations of HVA and 5-HIAA were measured by high-performance liquid chromatography equipped with electrochemical sensors, as previously described.^9^ Reference values of HVA and 5-HIAA were based on the results obtained from five controls without neurological diseases at autopsy.^9^ CSF HVA results from the patients taking antiparkinsonian drugs were excluded, as these medications are known to increase the level of CSF HVA.

### ApoE Phenotyping

Phenotyping of ApoE was performed by isoelectric focusing followed by Western blotting.

### Definition of Positive and Negative Aβ Accumulation in Amyloid PET

Those patients with a diagnosis of NIID who consented (n = 6) underwent [^11^C] Pittsburgh compound B (PiB) PET to assess the burden of Aβ, as described previously.^22^ Briefly, static emission data were acquired for 40–60 min after intravenous bolus injections of [^11^C] PiB. The dose was approximately 500 MBq. Amyloid status was visual determined by two experts (K. Ishibashi and K. Ishii), and amyloid positivity was defined as being not negative according to standard criteria.^23^ Aβ levels were quantified on the Centiloid (CL) scale using CapAIBL.^24^ A cutoff value of 12.2 CL has previously been reported for detecting Aβ depositions in the brain.^25^

### Genetic Analyses

One patient (Patient 4) was included in a previous report identifying *NOTCH2NLC* CGG repeat expansion in NIID.^14^ Three patients who presented to our hospital after the identification of *NOTCH2NLC* CGG repeat expansion in NIID were tested for this repeat expansion by repeat-primed PCR, as previously described at the University of Tokyo.^14^ Among those patients who were lost to follow-up before the identification of *NOTCH2NLC* CGC repeat expansion in NIID, three patients consented for genetic testing for *FMR1* GGC repeat expansion to rule out fragile X-associated tremor/ataxia syndrome (FXTAS) as a diagnostic workup. Repeat-primed PCR for *FMR1* GGC repeat expansion was conducted at Tottori University, as previously described.^26^

### Statistical Methods

Statistical analyses were conducted using the software programs R version 4.0.3 (R Foundation for Statistical Computing, Vienna, Austria), and a graphical interface EZR (Saitama Medical Center, Jichi Medical University, Saitama, Japan)^27^, or GraphPad Prism version 9 (GraphPad Software, San Diego, CA, USA). Missing data were handled using pairwise deletion approach. Categorical variables are presented as percentages, and differences between groups were evaluated using Fisher’s exact test. *Post hoc* analyses for multiple comparison were performed using Bonferroni correction. Continuous variables are presented as mean ± SD. Differences between groups were evaluated using Welch’s t-test or one-way analysis of variance (ANOVA) followed by *post hoc* analyses using Tukey’s test. Analysis of covariance (ANCOVA) was performed to evaluate the difference of CSF p-tau181 between NIID and disease controls using CSF t-tau as a covariate, after confirming the correlation between CSF p-tau181 and t-tau in each group. Pearson correlation analysis was conducted between CSF p-tau181 and other continuous variables in patients with NIID. Multivariate analysis was not conducted due to a limited number of NIID patients. P values of < 0.05 were considered statistically significant for all analyses.

## Results

### Clinical Features of the Patients with NIID

CSF results of 12 patients with NIID were available during the study period. Characteristics of each patient with NIID included in this study are summarized in **Table 1**. Three patients were male and nine patients were female. All patients were adult-onset, and initial symptoms were either dementia or paroxysmal symptoms, such as encephalitic attacks. The presence of CGG repeat expansion in *NOTCH2NLC* was confirmed in four patients, and GGC repeat expansion in *FMR1* was excluded in three of the remaining patients. At CSF collection, patients were aged 71.3 (mean) ± 4.4 (SD) years old, and the Mini-Mental State Examination (MMSE) and the Frontal Assessment Battery (FAB) scores were 20.6 ± 5.2 and 8.8 ± 3.0, respectively. Follow-up CSF collection was available in one patient immediately after an encephalitic attack four years after the first CSF collection (Patient 7). A summary of the characteristics of patients with NIID and disease controls is shown in **Table 2**.

**Table 1.**
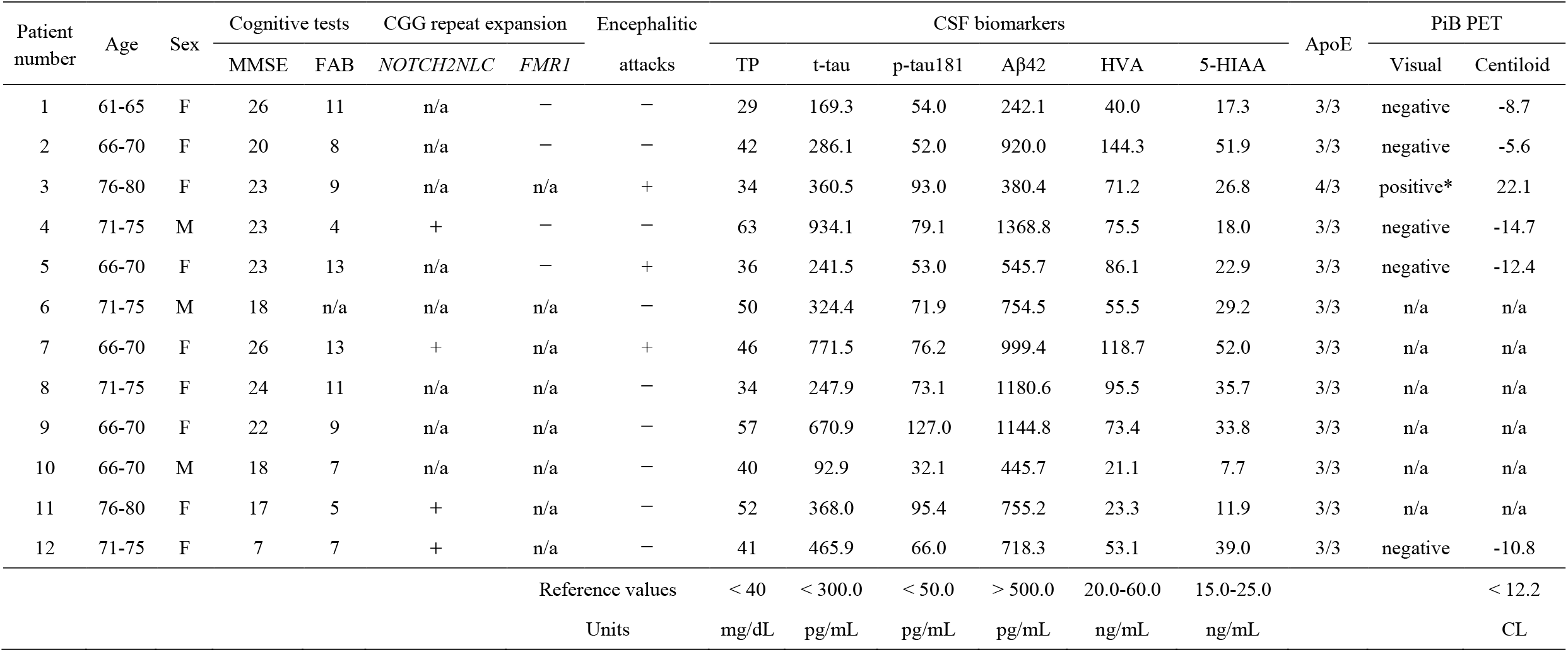
Characteristics and test results of each patient with NIID. Abbreviations: MMSE: Mini-Mental State Examination, FAB: Frontal Assessment Battery, TP: total protein, t-tau: total tau, p-tau181: tau phosphorylated at threonine 181, Aβ42: amyloid-beta 42, HVA: homovanillic acid, 5-HIAA: 5-hydroxyindole acetic acid, PiB: Pittsburgh compound B, CL: centiloid, n/a: not available, *: mild local accumulation

**Table 2.** Demographics of patients with NIID and disease controls. NIID: neuronal intranuclear inclusion disease, AD: Alzheimer’s disease, DLB: dementia with Lewy bodies, FTD: frontotemporal dementia, PSP: progressive supranuclear palsy, CBS: corticobasal syndrome.

|  | NIID<br>(n = 12) | AD<br>(n = 120) | DLB<br>(n = 24) | FTD<br>(n = 13) | PSP<br>(n = 21) | CBS<br>(n = 13) |
| --- | --- | --- | --- | --- | --- | --- |
| Age (years old) | 71.3 ± 4.4 | 74.6 ± 9.4 | 76.8 ± 8.4 | 70.2 ± 8.1 | 75.5 ± 5.2 | 71.9 ± 5.4 |
| Female (%) | 75.0% | 63.3% | 41.7% | 53.8% | 52.3% | 30.8% |
| ApoE4 carrier (%) | 8.3% | 50.4% | 40.9% | 8.3% | 25.0% | 0.0% |

### CSF p-tau181 Was Increased in Patients with NIID

CSF p-tau181 was significantly higher in NIID compared to DLB, PSP, and CBS (**Figure 1A**), and was comparable between NIID and AD (94.7 ± 33.6 vs 72.7 ± 24.8 pg/mL). CSF Aβ42 was decreased in AD, DLB, and PSP, and was significantly higher in NIID compared to AD, DLB, and PSP (**Figure 1B**). CSF t-tau was significantly higher in patients with AD, as compared to NIID, DLB, FTD, PSP, and CBS. CSF t-tau also showed a higher trend in NIID (**Figure 1C**). CSF p-tau181 and t-tau showed moderate correlation in NIID, AD, DLB, and CBS. The least-square regression line was similar between NIID and AD (**Figure 1D**), and the difference of CSF p-tau181 between NIID and AD was non-significant (p = 0.56) by ANCOVA, adjusted by CSF t-tau, while the difference between NIID and DLB, and that between NIID and CBS, was significant (p = 0.0024 and 0.0077, respectively).

**Figure 1.**
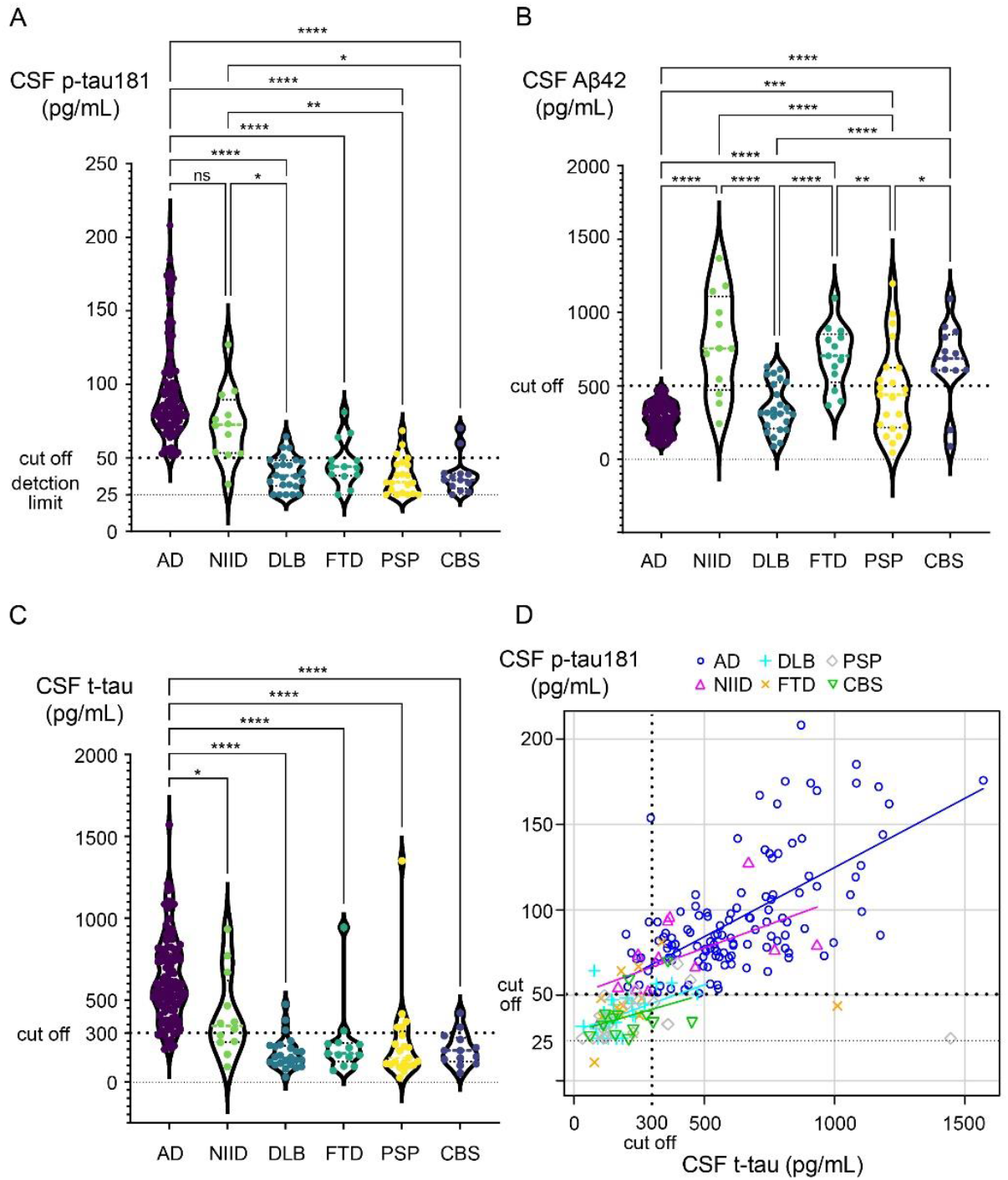
AD-related CSF biomarkers in patients with NIID and disease controls. (A) CSF p-tau181 was significantly higher in AD compared to DLB, FTD, PSP, and CBS, as previously reported. CSF p-tau181 was significantly higher in NIID compared to DLB, PSP, and CBS. (B) CSF Aβ42 was decreased in AD, DLB, and PSP. CSF Aβ42 was higher in NIID as compared to AD, DLB, and PSP, and was comparable to FTD and CBS. (C) CSF t-tau was significantly higher in AD as compared to NIID, DLB, FTD, PSP, and CBS, as previously reported. CSF t-tau also showed a higher trend in NIID, although non-significant. (D) CSF p-tau181 and t-tau showed moderate correlation in NIID (r = 0.57), AD (r = 0.62), DLB (r = 0.50), and CBS (r = 0.37). The difference of CSF p-tau181 between NIID and AD was non-significant by ANCOVA adjusted by CSF t-tau (p = 0.56).

### Increased CSF p-tau181 in Patients with NIID Was Not Related to Decreased CSF Aβ42

The results of CSF Aβ42 and p-tau181 of each patient are plotted in **Figure 2A**. Based on predetermined institutional cutoff values, quadrants were divided into the following categories: A-T-, A+T-, A-T+, or A+T+ (**Figure 2A**). The percentage of each category in each disease group is summarized in **Figure 2B**. While 75% of patients with NIID were classified as A-T+, only 4.2–23% were in non-AD disease controls and the frequency of A-T+ was significantly higher in NIID as compared to AD, DLB, PSP, and CBS (**Figure 2B**).

**Figure 2.**
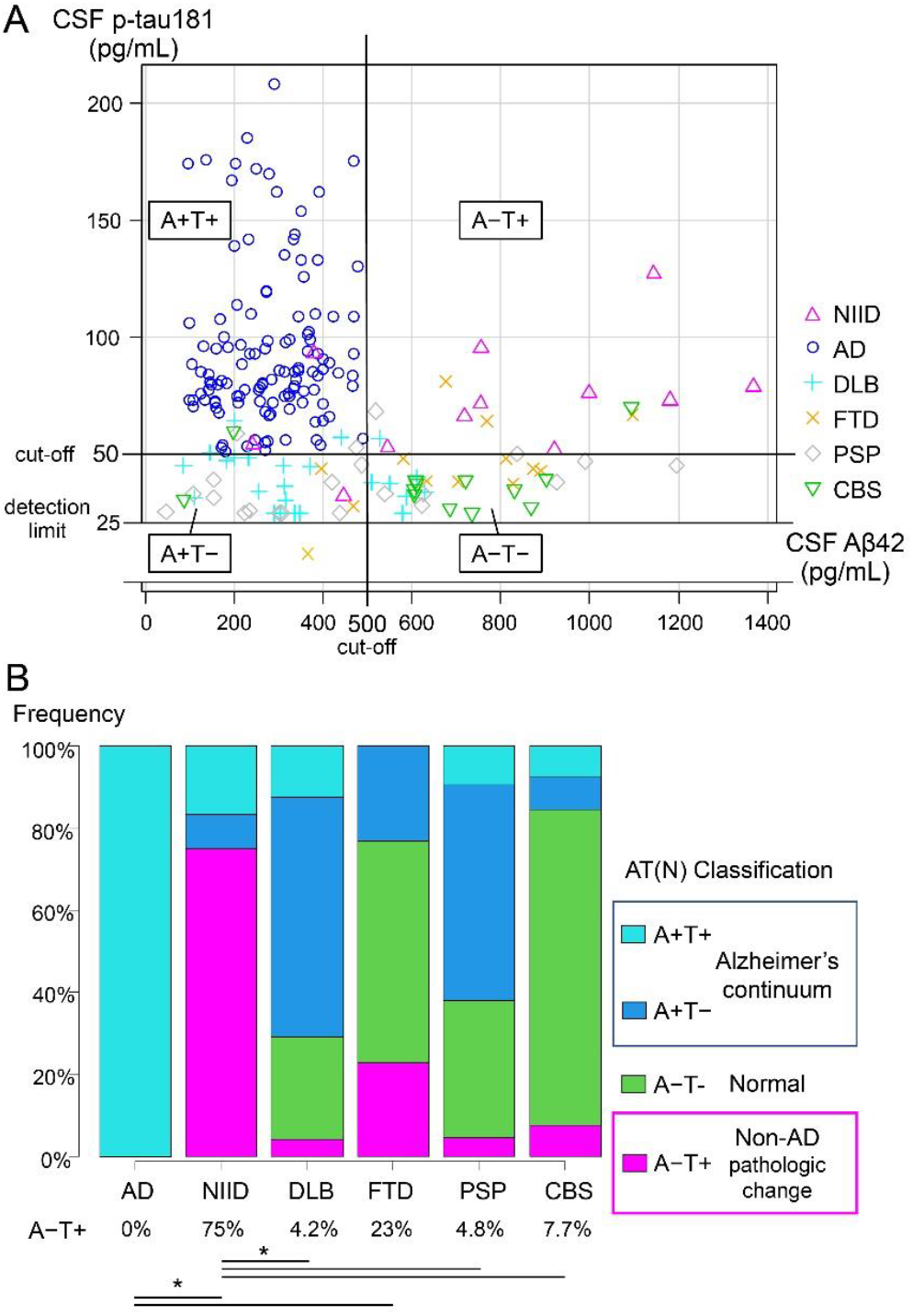
AT(N) classification in patients with NIID and disease controls. (A) Scatter plot of CSF Aβ42 and p-tau181 levels of each patient. Quadrants are divided into A-T-, A+T-, A-T+, or A+T+ according to the presence of abnormal CSF Aβ42 (A) and p-tau181 (T) results using predetermined institutional cutoffs. (B) Bar graph showing the frequency of each AT(N) classification in each disease groups. The frequency of A-T+ was significantly higher in NIID compared to AD, DLB, PSP, and CBS.

### PiB PET Imaging Showed Negative or Only Mild Local Accumulation in Patients with NIID

PiB PET imaging was conducted in six patients with NIID (patients 1–5 and 12), including both of the patients classified as A+T+ according to the CSF biomarker results (patients 1 and 3). Patient 3 showed mild local accumulation (22.1 CL), and the rest of the patients were amyloid negative by PiB PET (**Table 1**).

### CSF Concentrations of HVA and 5-HIAA Were Increased in Patients with NIID

The CSF concentrations of monoamine metabolites HVA and 5-HIAA were significantly higher in patients with NIID as compared to disease controls (**Figure 3A, B**). CSF concentrations of HVA and 5-HIAA in patients with DLB were lower than in patients with AD, as previously described (**Figure 3A, B**).^8,9^

**Figure 3.**
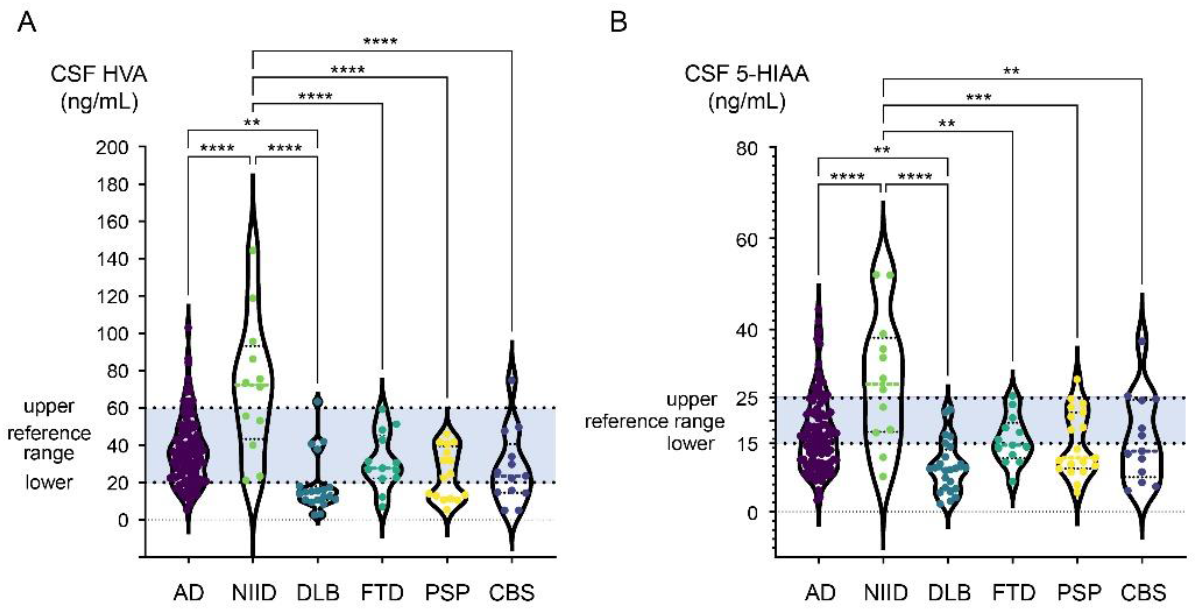
CSF monoamine metabolites in patients with NIID and disease controls. (A) CSF homovanillic acid (HVA) was significantly higher in NIID compared to AD, DLB, FTD, PSP, and CBS. CSF HVA was significantly lower in DLB compared to AD, as previously reported. (B) CSF 5-hydroxyindole acetic acid (5-HIAA) was significantly higher in NIID compared to AD, DLB, FTD, PSP, and CBS. CSF 5-HIAA was significantly lower in DLB as compared to AD, as previously reported.

### Factors Influencing CSF p-tau181 in Patients with NIID

Age at CSF, CSF total protein, CSF Aβ42, CSF HVA, CSF 5-HIAA, the MMSE score, and the FAB score did not show significant correlation with CSF p-tau181. No differences based on sex were observed (female 76.6 ± 24.9 vs male 61.0 ± 25.3 pg/mL).

### CSF Follow-up in One NIID Patient After Encephalitic Episode Showed No Change in CSF p-tau181 Level

Although encephalitis is known to cause transient CSF p-tau181 elevation^28,29^ and the subsets of NIID patients are known to experience encephalitic/stroke-like attacks,^10,11^ CSF p-tau181 levels of patients with or without a history of encephalitic/stroke-like attacks were also comparable (74.1 ± 20.1 vs 72.3 ± 27.3 pg/mL). In patient 7, CSF was available at baseline, during the remission phase, and one day after an encephalitic attack four years later as a follow-up. During the four years, the patient experienced mild cognitive decline and began to require assistance in preparing dinners, but remained independent in daily activities during remission. Her baseline and follow-up scores of the MMSE were 26 and 27 and of FAB were 13 and 11, respectively. All CSF parameters including p-tau181 were similar between baseline and follow-up (p-tau181, 76.2/77.7 pg/mL; Aβ42, 999.4/1062 pg/mL; t-tau, 771.5/628.7 pg/mL; HVA, 118.7/123.7 ng/mL; 5-HIAA, 52.0/59.0 ng/mL; respectively).

## Discussion

CSF p-tau181 was increased in patients with NIID even in the absence of biomarkers suggesting amyloid pathology. CSF concentrations of HVA and 5-HIAA were also higher in patients with NIID than in patients with other neurocognitive disorders.

Tau is a microtubule-associated protein and is mainly expressed in neuronal axons in the brain. It is widely known that CSF t-tau and p-tau181 are both increased in patients with AD.^4,30^ While t-tau can be increased in conditions with acute neuronal injury or degeneration such as stroke,^31^ head trauma,^32^ or Creutzfeldt–Jakob disease (CJD),^33^ consistent increase in p-tau181 has not been observed in any other neurodegenerative diseases, including primary tauopathies, and is considered specific to AD.^1,4,5^ CSF p-tau181 can even be decreased in CJD despite increased t-tau.^33^ Due to this specificity, increased CSF p-tau181 may even suggest AD co-pathology in patients with other neurodegenerative diseases, such as DLB.^9,34^ Because of the early increase and high specificity to AD, it has recently been recognized that increased CSF p-tau181 reflects neuronal secretion of p-tau associated with Aβ pathology in the brain.^5,6^

Therefore, we found it surprising that most patients with NIID showed increased CSF p-tau181 in the absence of biomarkers suggesting amyloid pathology in this study. NIID is characterized by eosinophilic hyaline intranuclear inclusions and is caused by CGG repeat expansions in *NOTCH2NLC*.^14,15^ Patients most often present with gradual cognitive decline, and brain MRI shows leukoencephalopathy with characteristic high intensity signal along the corticomedullary junction by diffusion-weighted imaging.^10^ Previous autopsy studies consistently reported that astrocytes are more affected than neurons in adult-onset NIID,^35,36^ and that the intranuclear inclusions are negative for Aβ or p-tau.^37–39^ No relationship between NIID and Aβ or p-tau has been suggested to date. There was a single case report of a patient with NIID showing markedly high CSF p-tau protein level of 167 pg/mL with normal CSF Aβ40/Aβ42 ratio and PiB PET.^40^ Although the authors speculated that an increase in CSF p-tau was most likely due to neuronal loss, the progression of the symptom was very slow,^40^ as compared to acute neurodestructive disorders known to cause CSF t-tau elevation, such as stroke, head trauma, and CJD.^31–33^ We showed that CSF p-tau181 was increased in most patients with NIID despite slow disease progression of this disease. Our CSF p-tau181/t-tau ratio of NIID comparable to AD also suggested that this increase was more likely due to a similar mechanism involved in AD (e.g., the secretion of p-tau from neurons through an unknown mechanism),^5,6^ rather than a non-specific increase due to acute neurodestruction in stroke, head trauma, or CJD.

Previous studies suggested that viral encephalitis was an important exception of CSF p-tau181 elevation.^28,29^ Krut et al. reported that patients with herpes-simplex virus type 1 (HSV-1) encephalitis showed increased CSF p-tau181 and t-tau along with decreased CSF Aβ42.^28^ As previous *in vitro* studies showed that the HSV-1 virus could induce hyperphosphorylation of tau,^41,42^ possibly through the activation of phosphokinases such as GSK3β,^42^ the authors suggested that an increase of CSF p-tau181 could be due to the direct effect of HSV-1.^28^ It would be intriguing to speculate that the phosphorylation of tau may be increased in patients with NIID. Although encephalitic attacks due to unknown mechanism are frequently observed in patients with NIID,^10,11^ our results suggested that an increase in CSF p-tau181 in patients with NIID was not a transient phenomenon related to these attacks.

It is reported that the phosphorylation of tau at threonine 181 can be caused by several phosphokinases, including GSK3β, Cdk5, and MAPKs.^43,44^ The pathophysiology of NIID is still being studied, and it is not yet known whether these kinases are altered in NIID. Recently, the translation of GGC repeat expansions into a toxic polyglycine protein similar to FXTAS has been reported in NIID using cellular and mouse models.^45,46^ These models or the patient-derived induced pluripotent stem cells (iPSCs)^47^ would definitely accelerate the pathophysiological research of NIID and may disclose the underlying mechanism of CSF p-tau181 increase in patients with NIID.

CSF concentrations of HVA and 5-HIAA are decreased in patients with DLB, likely reflecting reduced dopamine and serotonin in the brain, respectively.^8,9^ Surprisingly, CSF concentrations of HVA and 5-HIAA were higher in patients with NIID than in all the disease controls included in this study. HVA is metabolized from dopamine by monoaminoxidase (MAO) and catechol-O-methyltransferase, and 5-HIAA is metabolized from serotonin by monoaminoxidase.^7^ Our results suggest that the brain concentrations of dopamine and serotonin themselves or their metabolism, such as MAO activity, may be differentially altered in NIID and other neurocognitive disorders.

There are several limitations to this study. First, the number of patients with NIID was relatively small. Second, autopsy was not available to rule out tau deposition in the brain of the studied patients. Future studies are needed to elucidate the underlying mechanism behind the increase in CSF p-tau181 in patients with NIID. Third, we have not tested other promising CSF p-tau biomarkers reported to be more specific to AD, such as p-tau217 and p-tau231.^48–50^ It would be interesting to know whether these biomarkers are also increased in NIID.

In conclusion, CSF p-tau181 was increased in patients with NIID even in the absence of amyloid pathology. Although deposition of phosphorylated tau has not been reported in the brains of patients with NIID, molecular mechanism of tau phosphorylation or secretion of p-tau may be altered in NIID. CSF HVA and 5-HIAA was also elevated in patients with NIID. These results may be useful to understanding the pathophysiology of NIID.

## Data Availability

All data produced in the present study are available upon reasonable request to the authors

## Acknowledgements

We would like to thank all the patients and their families for participating in this study. We also thank members of the Department of Neuropathology (Brain Bank for Aging Research) and the Research Team for Neuroimaging at the Tokyo Metropolitan Institute of Gerontology for their technical assistance.

